# Antiherpetic medication and incident dementia: observational cohort studies in four countries

**DOI:** 10.1101/2020.12.03.20241497

**Authors:** Christian Schnier, Janet Janbek, Linda Williams, Tim Wilkinson, Thomas Munk Laursen, Gunhild Waldemar, Hartmut Richter, Karel Kostev, Richard Lathe, Jürgen Haas

## Abstract

**Introduction:** Recent meta-analysis of the association between herpesvirus infection and dementia concluded that the evidence for an association to date is insufficient.

**Methods:** 2.5 million individuals aged ≥65 years were followed up using linked electronic health records in four national observational cohort studies. Exposure and outcome were classified using coded data from primary and secondary care. Data were analyzed using survival analysis with time-dependent covariates.

**Results:** Results were heterogeneous, with a tendency towards decreased dementia risk in individuals exposed to antiherpetic medication. Associations were not affected by treatment number, herpes subtype, dementia subtype, or specific medication. In one cohort, individuals diagnosed with herpes but not exposed to antiherpetic medication were at higher dementia risk.

**Discussion:** Short-term antiherpetic medication is not markedly associated with incident dementia. Because neither dementia subtype nor herpes subtype modified the association, the small but significant decrease in dementia incidence with antiherpetic adminstration may reflect confounding and misclassification.

## 1. INTRODUCTION

The role of infections in the development of dementia has been widely debated. Attention has focused on herpes viruses infection in part because variants at the *APOE* locus are not only associated with Alzheimer’s disease (AD) (e.g., [1]) but also govern infection with herpes viruses (e.g., [2]). Moreover, infections with herpes simplex virus (HSV) and the related varicella zoster virus (VZV) are known to induce deposition of amyloid-β [3]–[5].

Several epidemiological studies from Taiwan, all using the same data resource, found significant associations between herpes virus infection and dementia. First, individuals diagnosed with herpes zoster ophtalmicus (caused by VZV) had an increased (HR = 3) rate of incident dementia within 5 years of diagnosis [6]; information on antiherpetic medication was not available. Second, patients with general VZV infection had a slightly increased risk of incident dementia (HR = 1.1) compared to controls [7], and the subgroup who were treated with antiherpetic medication had a reduced risk of dementia (HR = 0.6) compared to untreated HSV-positive patients. Third, HSV infection was reported to increase the risk of incident dementia (HR = 2.5) [8], and treatment with antiherpetic medication markedly reduced the risk (HR = 0.1 versus untreated HSV-positive controls). Moreover, the association was greater in those treated for longer times. In addition, a study from South Korea reported that patients with VZV infection had an increased rate of incident dementia (HR = 1.1), but this was reduced (HR = 0.8 versus untreated VZV) by antiherpetic medications [9].

Despite these reports, a recent meta-analysis into associations of human herpesvirus infections with dementia or mild cognitive impairment concluded that evidence to date for an association is insufficient [10].

We therefore conducted a multicenter observational cohort study using health registry data from Wales, Germany, Scotland, and Denmark to investigate potential associations between antiherpetic medication and incident dementia, and also to comprehensively investigate such associations broken down according to medication type and dose, type of herpes virus, and dementia subtype.

## 2. METHODS

The primary objective was to study the association between oral antiherpetic medication and incident dementia. Secondary objectives were to study whether that association was modified by (i) number of treatments; (ii) herpes subtype; (iii) type of antiherpetic medication, and (iv) dementia subtype.

### 2.1 Data sources

We utilized linked routinely collected health data (RCHD) from four different sources: (i) the Secure Anonymised Information Linkage Databank (SAIL), Wales; (ii) the IMS® Disease Analyzer, Germany; (iii) the Danish National Registries (DNR); and (iv) the Electronic Data Research and Innovation Service (eDRIS) of Public Health, Scotland.

The SAIL Databank holds anonymized health data from ∼80% of Welsh general practioner practices linked to national datasets including inpatient records, death records, and Welsh Index of Multiple Deprivation (WIMD) [11]. The IMS Disease Analyzer database holds unlinked and anonymized information on drug prescriptions, diagnoses, and basic medical and demographic data from ∼3% of all outpatient medical practices in Germany [12]. The DNR comprise linked RCHD from the entire Danish population; registers used in this study were the Danish Civil Registration System[13], the Danish National Patient Registry[14], and the Danish National Prescription Registry[15]. The eDRIS database holds countrywide linked anonymous RCHD on hospital admissions, prescriptions, mortality, and national health service registration data [16]. All four datasets are representative of the general population in the relevant country; however, individuals with poor quality/missing identifier data or reduced access to healthcare (e.g., migrants) are likely to be under-represented.

### 2.2 Study populations

The study populations in SAIL, IMS Disease Analyzer, and eDRIS comprised all individuals with follow-up data from age 60 onwards who had no dementia-related record before their 65th birthday, were alive at their 65th birthday, and for whom no information on any of the covariates in the analysis was missing (Figure 1). The study population in DNR comprised all individuals born in or before 1950, alive and residing in Denmark, with no dementia-related RCHD before start of follow-up. In IMS Disease Analyzer, individuals were excluded for whom a dementia-related prescription but no dementia-related diagnosis could be found or who received antiherpetic medication during follow-up without any known herpes-related diagnosis. Start of follow-up in cohorts from SAIL, IMS Disease Analyzer, and eDRIS was the 65th birthday (mid-month in datasets where only month and year of birth were available, or mid-year where only year of birth was available). In the DNR cohort, start of follow-up was the earlier of 1 January 2000 and 65th birthday. End of follow-up was at the date of the first dementia-related code or the date of deregistration, death, or the end of the study period. In the unlinked data from IMS Disease Analyzer, mortality data were not available; however, a death would have been recorded as deregistration (Table 1).

**Table 1.**
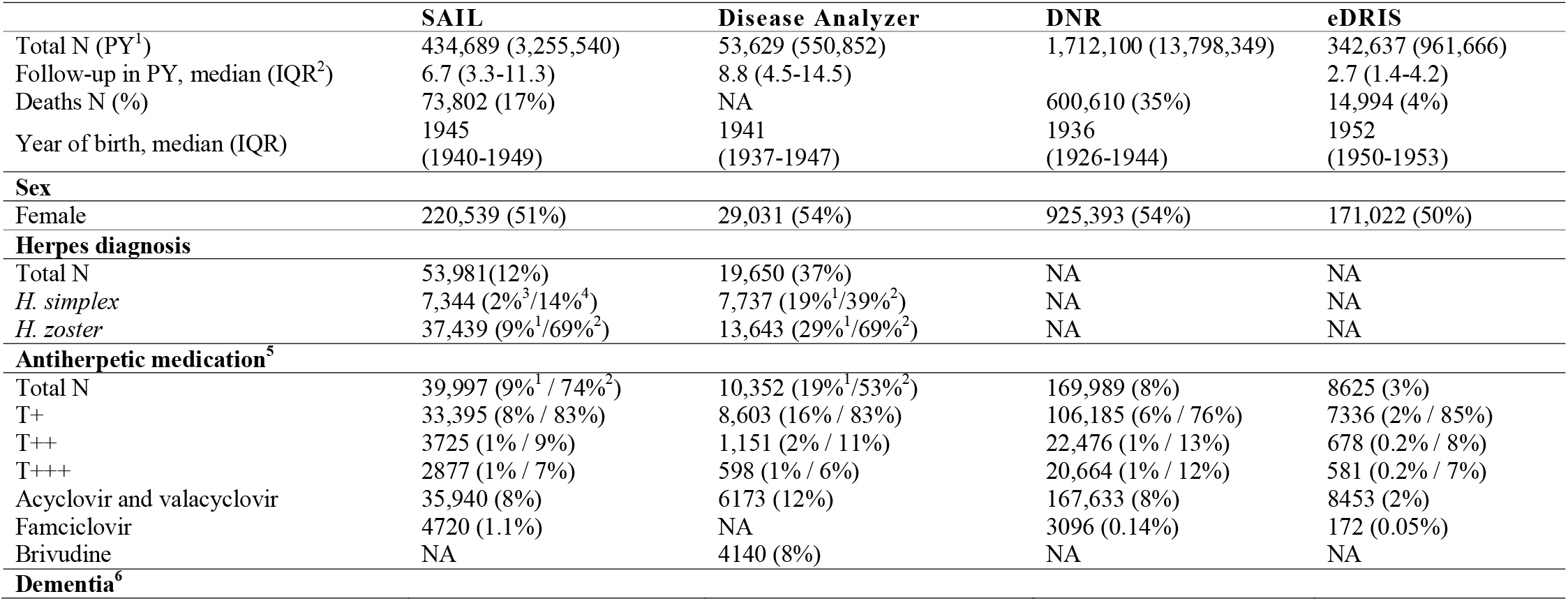
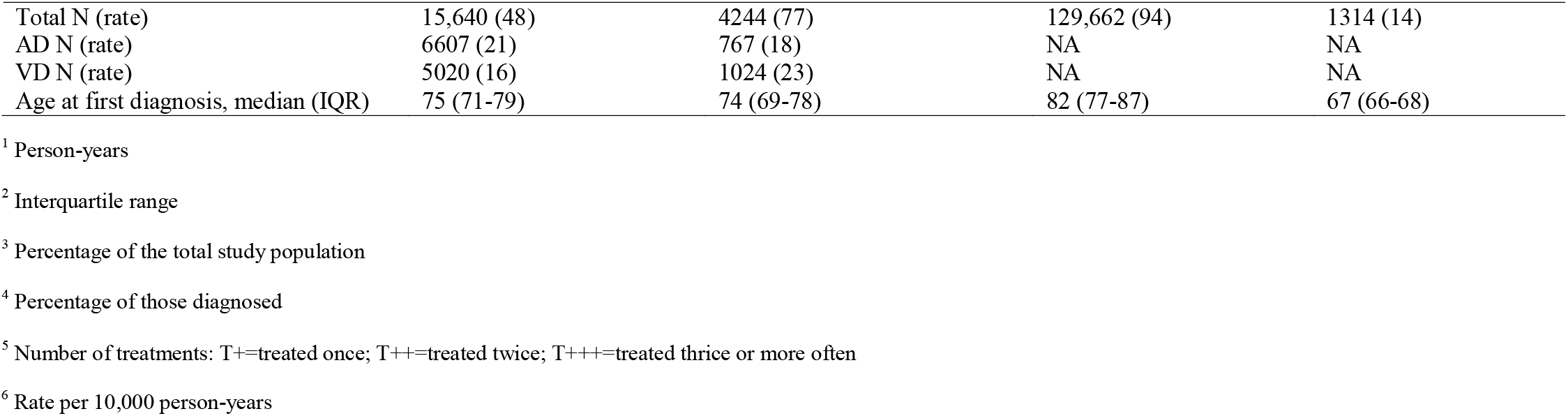
Study population descriptive data.

**Figure 1.**
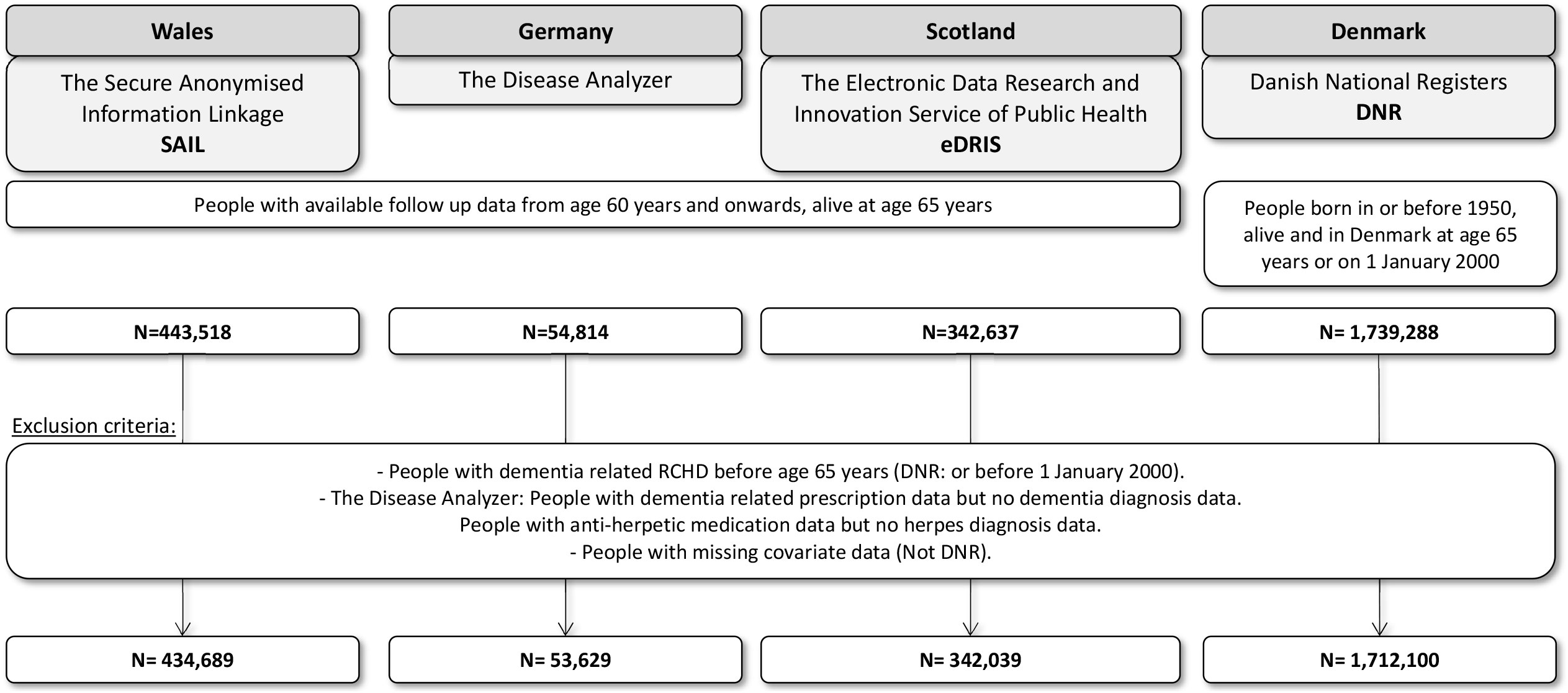
Flow chart of the four study populations.

### 2.3. Dementia classification

In all datasets we used coded information to classify individuals with dementia, using the date of the first code as date of diagnosis. Table S1 lists codes used for dementia classification. In SAIL, individuals were classified as having dementia if they had a dementia-related diagnostic code from primary care, hospital admission, or mortality records (underlying or contributing cause). In the unlinked data from IMS Disease Analyzer, individuals were classified as having dementia if a dementia-related diagnosis was documented from primary care data. In DNR and eDRIS, individuals were classified as having dementia if they had a dementia-related prescription or dementia-related hospitalization. In cohorts with available diagnostic codes from primary care (SAIL and IMS Disease Analyzer), diagnostic codes were also used to subtype dementia into AD, vascular dementia (VD), and ‘other/unknown’. Individuals with codes for both subtypes were counted as both AD and VD (‘mixed dementia’ is not coded in the RCHD). The positive predictive value (PPV) of RCHD (diagnostic codes from primary care, hospitalization, and mortality records) to classify dementia and dementia subtypes has recently been studied in Scotland, and the data showed good PPV for classification of dementia and AD, but relatively low PPV for classifying VD [17].

### 2.4 Antiherpetic drugs

In all datasets, exposure to antiherpetic medication was restricted to oral antiherpetic medication; topical applications can be purchased without prescription and are therefore under-reported in RCHD. Exposure classification was also restricted to prescriptions from primary care because those from hospital admissions were not available. In all four countries prescriptions from primary or secondary care are necessary for access to oral antiherpetic medication; however, medication can be prescribed over the internet, in which case exposure would not be recorded in our databases. Table S1 lists codes used for antiherpetic medication classification.

### 2.5 Herpes classification

In SAIL and IMS Disease Analyzer, codes from primary care diagnosis and hospital admissions were used to classify herpes infection and herpes subtypes (*Herpes simplex* and *H. zoster*), and the date of the first diagnostic code was taken as the date of diagnosis. Individuals with diagnostic codes for both subtypes were classified according to the diagnostic code entered at first diagnosis (diagnostic codes in Table S1).

### 2.6 Covariates

Linked RCHD were used to classify year of birth and sex. We used several proxies for socioeconomic status (SES): in SAIL and eDRIS we adjusted for multiple deprivation; in IMS Disease Analyzer we adjusted for type of insurance (private insurance being indicative of higher disposable income); and in DNR we adjusted for highest education and civil status at start of follow-up. In DNR, we additionally adjusted for comorbidities using the Charlson comorbidity index (excluding dementia).

### 2.7 Type of herpes infection

To investigate in SAIL and IMS Disease Analyzer whether the association of antiherpetic medication with incident dementia was affected by herpes subtype we conducted a subset analysis that included only individuals with either *H. zoster* infection or without any herpes diagnosis; individuals with a diagnosis of *H. simplex* were excluded. We then repeated the analysis including only individuals with *H. simplex* diagnosis and removed those with *H. zoster* infection. To study the effect of herpes subtype and associated treatment on dementia incidence in DNR and eDRIS, individuals were classified by the first prescribed drug (only aciclovir) into those with high drug strength (800 mg) and low drug strength (200 or 400 mg) that are indicative of *H. zoster* and *H. simplex* infection according to national prescribing guidelines in the two countries.

### 2.8 Type of dementia

To investigate in SAIL and IMS Disease Analyzer whether the association of antiherpetic medication with incident dementia was only seen in a particular type of dementia, we again explored a subset of the study population which included only individuals with either a diagnosis of AD or without any dementia, and excluded individuals with any other or ‘unknown’ dementia. We then repeated the analysis with VD.

### 2.9 Type of antiherpetic medication and number of treatments

For medication-specific analysis we included aciclovir and its prodrug, valaciclovir (combined), famciclovir (not IMS Disease Analyzer), and brivudine (only IMS Disease Analyzer). The original analysis included ganciclovir and valganciclovir; however, we excluded them because very few individuals in each cohort received these medications. In eDRIS, from the entire cohort of elderly individuals, only 105 received any antiherpetic other than aciclovir or valaciclovir, threfore medication-specific analysis was not performed. To study the effect of type of antiherpetic medication in SAIL and IMS Disease Analyzer we tested the effect of each drug by excluding individuals from the study population who were exclusively exposed to another antiherpetic medication. The number of prescriptions (one, two, ≥3) was analysed independently of the time between prescriptions.

### 2.10 Statistical analysis

Data analysis employed multivariable survival analysis. In all cohorts, sequential prescriptions of antiherpetic medication were included as time-dependent variables, increasing from zero prescriptions (unexposed controls) to ≥3 prescriptions. For cohorts with information on primary care diagnosis (SAIL and IMS Disease Analyzer), we additionally included herpes diagnosis as a time-dependent variable. In DNR, we used Poisson regression to fit piecewise exponential survival models with the logarithm of person years at risk as an offset variable [18](SAS software, Version 9.4 of the SAS System for Windows). To control for effects of age and year in these models, incremental years were included as time-dependent variables. In all other cohorts, we used Cox proportional hazard models (R, package survival [19]). To control for the effect of calendar year we adjusted for birth-year categories; the effects of age were accounted for by following up every individual in the cohort from their 65th birthday. For cohorts with information from primary care we included the practice number as random effect to control for correlation between patients of the same practice (R package coxme[20]).

### 2.11 Ethics and Governance

Analysis of data from SAIL and eDRIS was granted under IGRP 0938 and PBPP 1819-0297, respectively. Analysis of data from IMS Disease Analyzer and DNR did not require specific governance approval.

## 3. RESULTS

### 3.1 Study populations and drug prescription

There was considerable heterogeneity in the composition of the four study populations (Table 1), notably in mean follow-up time, ranging from 2.7 years (eDRIS) to 9 years (IMS Disease Analyzer). The longer follow-up time was associated with higher proportions of individuals being recorded as receiving antiherpetic medication and diagnoses of herpes virus infection and dementia. Similarly, including individuals with a start date later than their 65th birthday in DNR increased the average age of the DNR cohort and increased the rate of dementia diagnosis and death. The proportion diagnosed with herpes viral infection was higher in IMS Disease Analyzer than in SAIL, and *H. zoster* was diagnosed more often than *H. simplex* in both cohorts. In all four cohorts, aciclovir and valaciclovir were the most commonly prescribed antiherpetics; however, in IMS Disease Analyzer many prescriptions were for brivudine, a drug that was not prescribed in any of the other cohorts. In all four cohorts, antiherpetic medication was most commonly prescribed only once during follow-up, and at most 1% of all individuals in the cohorts were exposed to three or more doses of any antiherpetic medication. In both cohorts where dementia could be subtyped (SAIL and IMS Disease Analyzer) the number of individuals with dementia coded as AD was similar to the number coded as VD. In all cohorts, women were more likely to be exposed to antiherpetic medication than men (Table S2).

### 3.2 Dementia incidence

In all cohorts, exposure to one or more doses of antiherpetic medication was either significantly associated with a slight decrease in dementia incidence (adjusted hazard ratios ranged from HR = 0.89; 95% CI: 0.83–0.95; to HR = 0.93; 95% CI: 0.88–0.98) or was not significantly associated (Figure 2 and Tables S3–S6). Notably, in DNR (the largest cohort included in our study) all levels of exposure were associated with a small but significantly reduced incidence of dementia. Increased treatment numbers were not associated with differences in effects of exposure in any of the cohorts. Adjustment for covariates in the different cohorts did not have a large effect on any of the estimated HRs (Tables S3–S6). In IMS Disease Analyzer, individuals diagnosed with herpes virus infection but not exposed to any antiherpetic medication had an increased incidence of dementia compared to those not diagnosed (HR = 1.18; 95% CI: 1.09–1.28) (Figure 2 and Table S4). This increased rate was not seen in the other cohort that had information on diagnosis (SAIL) where individuals diagnosed but not treated had a non-significant lower incidence (HR = 0.95; 95% CI: 0.88– 1.02) (Figure 2 and Table S3).

**Figure 2.**
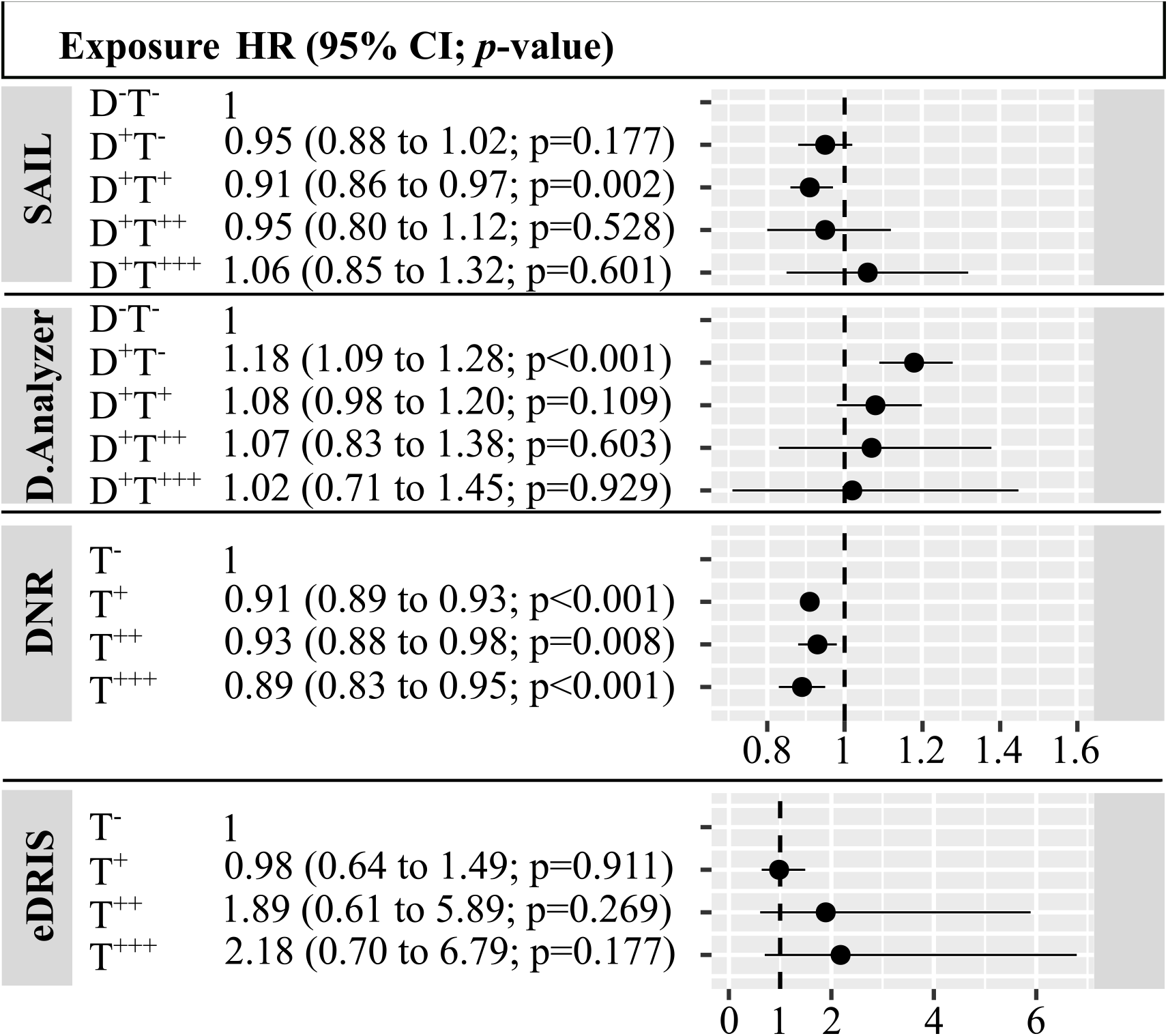
Association of exposure to antiherpetic medication with dementia incidence in four national cohorts. D-T-=not diagnosed, not treated; D+T- = diagnosed, not treated; D+T+ = diagnosed and treated once; D+T++ = diagnosed and treated twice; D+T+++= diagnosed and treated thrice or more often. Note the different scale of the x-axis for eDRIS compared to the other cohorts.

In all cohorts, effects of exposure to antiherpetic medications was independent of medication type (Figure 3). In IMS Disease Analyzer individuals diagnosed with *H. zoster* and not exposed to any antiherpetic medication had a higher rate of dementia (HR = 1.2; 95% CI 1.06–1.35) (Figure 4) compared to those undiagnosed, whereas dementia incidence in those diagnosed with *H. simplex* and not exposed was unchanged (HR = 0.97; 95% CI 0.88–1.07) (Figure 4). This trend towards higher HRs in individuals diagnosed with *H. zoster*, and lower HRs in those diagnosed with *H. simplex*, could also be observed in people exposed to antiherpetic medication; however, these differences were not significant (Figure 4). In SAIL and DNR, herpes subtype had no effect on the association between exposure to antiherpetic medication and dementia (Figure 4). Finally, in both SAIL and IMS Disease Analyzer the association between exposure to antiherpetic medication and dementia incidence was similar across dementia subtypes (Figure 5 and Tables S4,S5).

**Figure 3.**
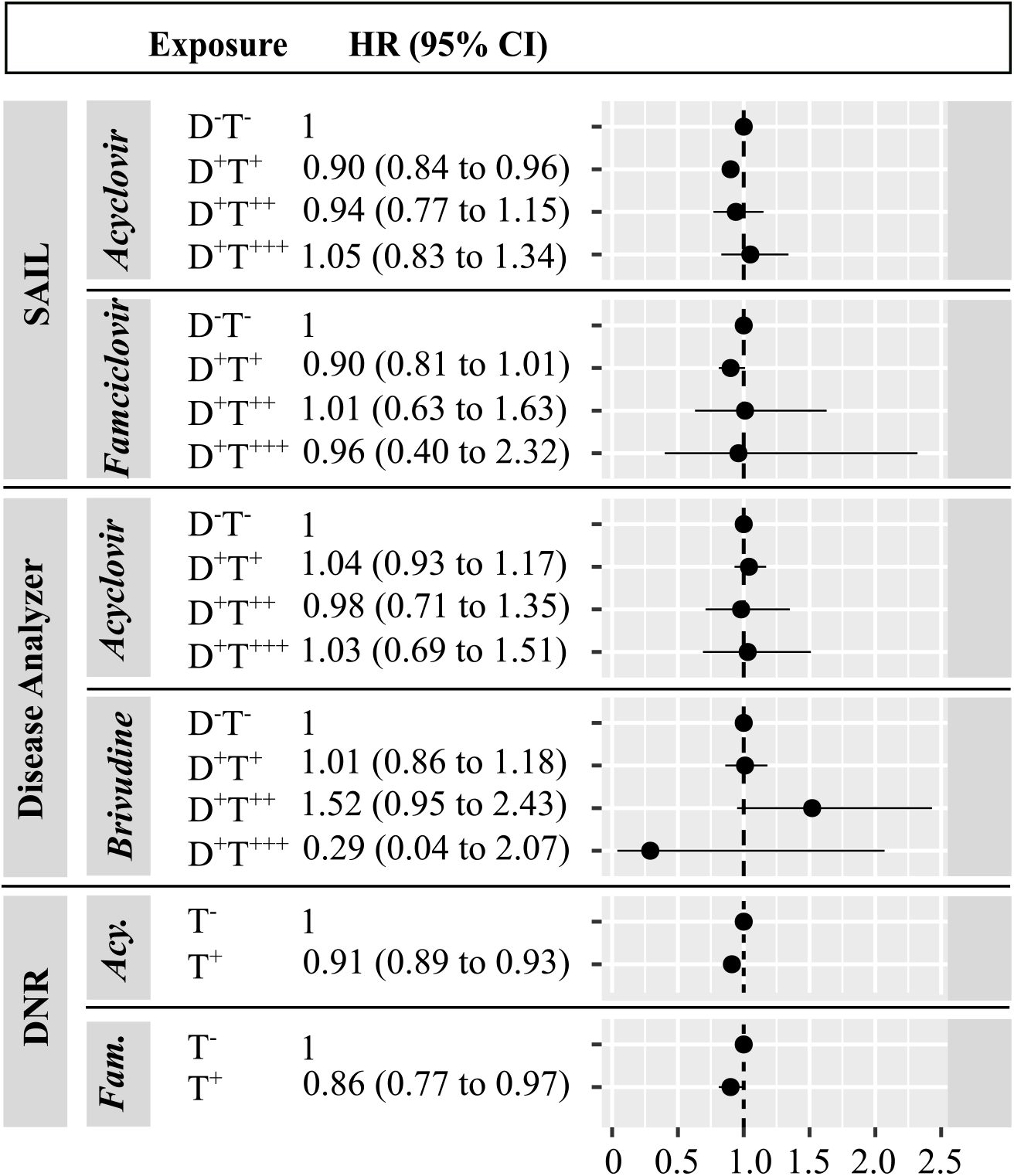
Association of exposure to antiherpetic medication with dementia incidence by type of antiherpetic medication. D-T-=not diagnosed, not treated; D+T+ = diagnosed and treated once; D+T++ = diagnosed and treated twice; D+T+++= diagnosed and treated thrice or more often.

**Figure 4.**
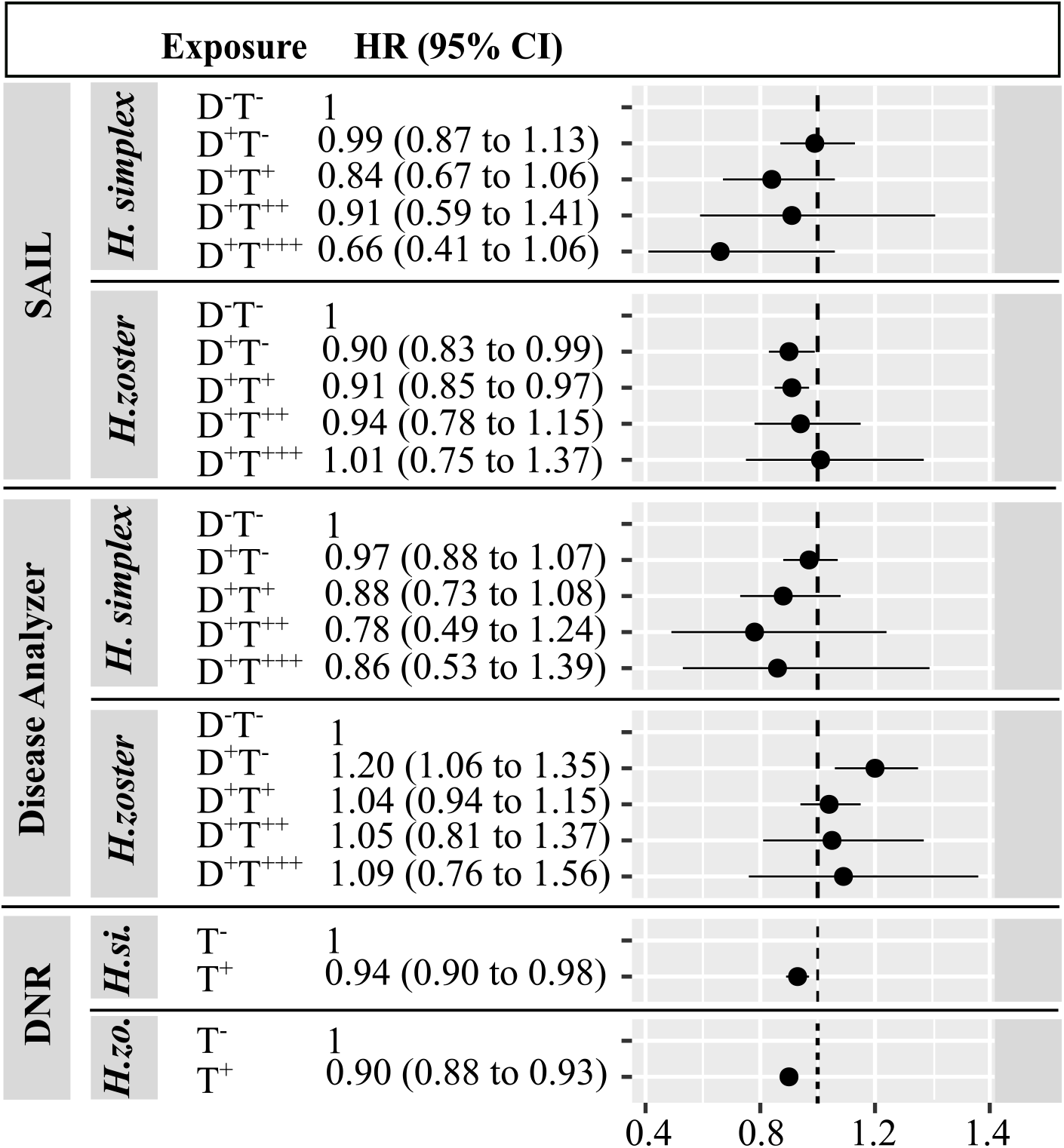
Association of exposure to antiherpetic medication with dementia incidence by underlying herpes infection. D-T-=not diagnosed, not treated; D+T- = diagnosed, not treated; D+T+ = diagnosed and treated once; D+T++ = diagnosed and treated twice; D+T+++= diagnosed and treated thrice or more often.

**Figure 5.**
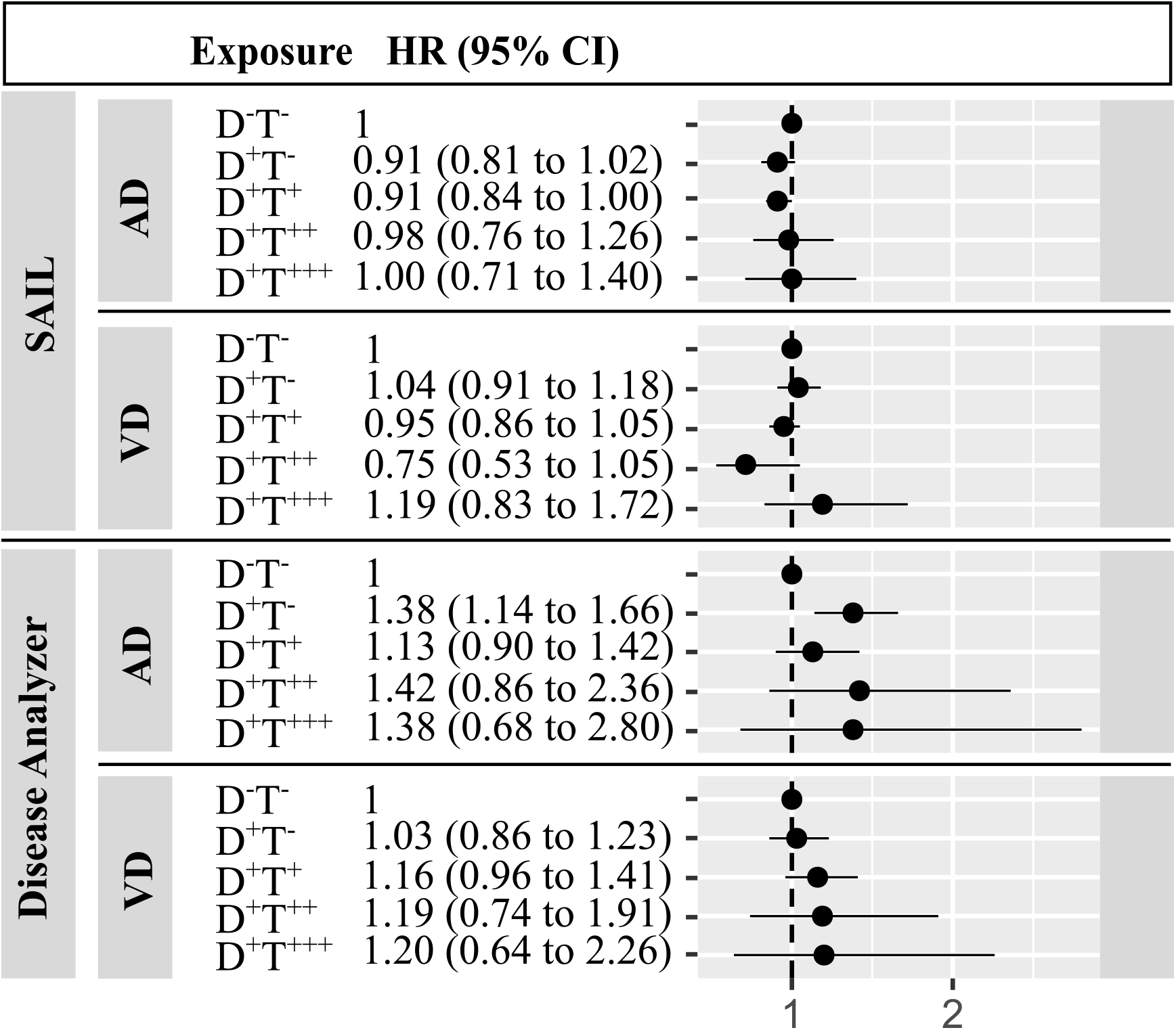
Association of exposure to antiherpetic medication with dementia incidence by type of dementia. D-T-=not diagnosed, not treated; D+T- = diagnosed, not treated; D+T+ = diagnosed and treated once; D+T++ = diagnosed and treated twice; D+T+++= diagnosed and treated thrice or more often.

## DISCUSSION

We analyzed associations between antiherpetic medication and dementia incidence in four large national observational cohorts. We report evidence for a low negative association between exposure and outcome; however, results were heterogeneous.

The negative association could be related to genuine protective effects of antiherpetic medication against neuropathologic consequences of herpes infection. Indeed, in IMS Disease Analyzer, individuals diagnosed with herpes infection but not treated were at higher risk of dementia compared to both those undiagnosed and those diagnosed and treated. This result resembles observations from Taiwan and South Korea [6], [8], [9]. However, there is evidence against this interpretation. First, the effect could not be seen in SAIL, the only other cohort with available diagnostic information. Second, the associations between antiherpetic medication and dementia in SAIL and IMS Disease Analyzer were for both AD and VD, diseases that have different neuropathological manifestations. Finally, the association was independent of the herpes subtype (*H. simplex* versus *H. zoster*), whereas possible associations of herpes infection with dementia have mostly been described for *H. simplex* and less for *H. zoster* [10].

Given that in cohorts other than IMS Disease Analyzer we did not see a dose– response effect, a direct neuroprotective effect of the antiherpetic medication seems unlikely. However, the great majority of patients in all four cohorts received single prescriptions of antiherpetic medications (typically for administration over a period of 1–2 weeks), and we cannot exclude the possibility that longer-term exposure might have greater effects.

The small negative association could also be related to indirect effects of exposure to antiherpetic medication. Given the nature of the observational cohorts, we could not study whether associations were affected by *APOE* genotype. Indeed, because a specific variant at the *APOE* locus, ε*4* (*APOE4*), is associated with susceptibility to both dementia and herpes virus infection, a positive association between dementia, herpes infection and antiherpetic medication would be expected. Of note, the Danish population in general has a slightly higher proportion of *APOE4* carriers than the other populations [21], which might have contributed to the observed heterogeneity.

Finally, the observed negative association could reflect residual confounding, reverse causation, and misclassification. There is clear evidence in all our study populations of a negative association between socioeconomic status (SES) and dementia incidence. SES could also be correlated with access to antiherpetic medication and medical care [22]; however, adjusting for SES in the statistical models did not change the estimated HR (Tables S3–S6). Similarly, controlling for the effects of comorbidities in DNR did not modify the significant negative association between exposure and dementia incidence (Table S5). Vaccination against VZV (shingles vaccine) has been reported to be associated with reduced dementia incidence [23]. However, the major effect of VZV vaccination would be to reduce herpes diagnosis and thereby exposure to antiherpetic medication, and therefore vaccination would not act as a confounder in our study. If preclinical dementia were to be associated with an increased risk of herpes diagnosis and antiherpetic medication, we would have falsely attributed the dementia diagnosis to the antiherpetic medication (reverse causation). This would mean that the true negative association was slightly larger than the observed HR. However, only <5% of all first antiherpetic medications were prescribed <6 months before dementia diagnosis. Finally, individuals in all cohorts might have been misclassified as unmedicated if they had received antiherpetic medication purchased outside the national public health system. However, our study population was restricted to the elderly population (65+), and we feel that exposure misclassification is less likely to have been significant.

Some of the heterogeneity could be explained by differences in study design and study populations. In DNR and eDRIS the non-exposed cohorts included diagnosed but untreated individuals; in the other cohorts these were included as a separate group. Including a mixture of diagnosed and untreated individuals and those undiagnosed and untreated could mask a true negative association if the association between herpes infection and incident dementia holds true: the risk of dementia would be unequal in the exposed and unexposed groups in these cohorts (DNR and eDRIS) in our study. By contrast, we have no explanation for the (non-significant) trend towards increased dementia risk in individuals exposed to antiherpetic medication in IMS Disease Analyzer. Interestingly, the proportion of individuals diagnosed with herpes as well as the proportion who were treated was much higher in IMS Disease Analyzer than in SAIL, even though individuals in the two cohorts were followed up for similar periods. Although brivudine was only used in IMS Disease Analyzer, the effects of brivudine exposure were similar to those of aciclovir/valaciclovir, and it seems unlikely that the type of antiherpetic medication might explain the heterogeneity. Heterogeneity in study design and populations could also explain the differences between our results and those from studies from Taiwan and South Korea where many individuals were exposed to substantially longer durations of medication and/or dosages.

Studies on dementia etiology require large study populations that are followed up over long periods of later life. Our study benefitted from the high quality of RCHD in the four nations containing reliable information on exposure, outcome, and covariates for a total of >2.5 million individuals aged >65 years. The size of the study populations allowed us to control for the single most important risk factor for dementia, age, by using a common age at the start of follow-up (IMS Disease Analyzer, SAIL, and eDRIS) or by using age as a time-dependent variable (DNR); in addition, we were able to include both repeated exposure to antiherpetic medication and herpes diagnosis as time-dependent covariates. However, our study has some limitations. Disease and exposure classification were based on imperfect information, especially regarding the validity of dementia subtypes, and the classification of herpes subtypes using drug strength as a proxy (in DNR). However, we would not expect misclassification to be differential. More importantly, because only information on the dates of dementia diagnosis and/or first dementia drug exposure were available, we could not ascertain whether exposure preceded or followed dementia development. Indeed, in the case of AD the pathology is likely to be present >10 years before symptom onset, so follow up time in our cohorts would have been insufficient. We could have excluded individuals with exposures to antiherpetic medication some period prior to dementia diagnosis; nevertheless, any time period would have been arbitrary and follow-up times in individuals without dementia impossible to calculate.

In conclusion, results from four large national cohorts indicate that short-term antiherpetic medication is not markedly associated with reduced dementia incidence. Because neither type of dementia nor type of herpes infection modified the association, the small but significant decrease in dementia incidence with antiherpetic administration may reflect unmeasured confounding and misclassification.

## Data Availability

Access to the underlying routinely collected electronic health records is governed by the different data provider (SAIL, eDRIS, IQVIA (epidemiology) and the Danish national research institute).

## Funding

We gratefully acknowledge financial support (grant VIRADE 2019) from the Benter Foundation, Pittsburgh, USA

## Notes

### Competing Interest Statement

Karel Kostev and Hartmut Richter are employees of IQVIA (Germany)

