## Supplemental tables S1 to S6 for "Antiherpetic medication and incident dementia: observational cohort studies in four countries"

**Supplementary material**

Table S1 Codes in RCHR used for classification of exposure and outcome.

Table S2 Study population characteristics by exposure status.

Table S3 Partially and fully adjusted results from the survival analysis model for the SAIL population.

Table S4 Partially and fully adjusted results from the survival analysis model for the IMS Disease Analyzer population.

Table S5 Partially and fully adjusted results from the survival analysis model for the Danish national registries population.

Table S6 Partially and fully adjusted results from the survival analysis model for the eDRIS population.

|  |  | **Dementia & dementia medication^[1]^** | | | **Herpes & antiherpetic medication** |
| --- | --- | --- | --- | --- | --- |
| **SAIL** | Primary care (READ V2^[2]^) | ^1461; ^38C13; ^3AE[3-6]; ^66h; ^6AB; ^8BM02; ^8CMG2; ^8CMZ[.0-3]; ^8CSA.; ^8Hla; ^8Iae2; ^9hD[.01]; ^9Ou; ^A411[.0]; ^E00[.0-4]; ^E012; ^E02y1; ^E041; ^Eu00[.012z]; ^Eu01[.0-3yz]; ^Eu02[.0-5yz]; ^Eu041; ^Eu10[67]; ^F110[.01]; ^F11[1268]; ^F11x[279]; ^F11y2; ^F21y2; ^ Fyu30 | | | ^A5[34] |
|  | Hospital admission (ICD 10) | ^F0[0-3]; ^F051; ^G30; ^G31[018]; ^I673 | | | ICD10: ^A60[019]; ^B0[02]; ^H191 |
|  | Prescription (READ V2) |  | | | ^ei[179CW] |
| **IMS Disease Analyzer** | Primary care (ICD 10) | ^F0[0-3]; ^G30; ^G31[018]; ^I673 | | | ^A60[019]; ^B0[02]; ^H191 |
|  | Prescription (EphMRA ATC^[3]^) | | ^N07D | | ^J05B3 |
| **DNR** | Hospital admission (ICD 8 and ICD 10) | | ^F0[0-3]; ^G30; ^G31[89]  ^290 | ^A60[019]; ^B0[02]; ^H191 | |
|  | Prescription (WHO ATC^[4]^) | | ^N04BA0[0-6]; ^N04BC0[459] | ^J05AB0[19]; ^J05AB11 | |
| **eDRIS** | Hospital admission (ICD 10) | | ^F0[0-3]; ^F051; ^G30; ^G31[018]; ^I673; ICD9: ^290; ^331[0-2] | ^A60[019]; ^B0[02]; ^H191 | |
|  | Prescription (BNF^[5]^) | | 4.11 | 5.3.2 | |

| **Source** | **Variable** | **Level** | **Exposure status** | |
| --- | --- | --- | --- | --- |
|  |  |  | **Antiherpetic medication** | **No antiherpetic medication** |
| **SAIL** | **Sex** | Male | 15,736 (39%) | 198,414 (50%) |
|  |  | Female | 24,261 (61%) | 196,278 (50%) |
|  | **WIMD^[6]^** | 1 (most deprived) | 6429 (16%) | 69,455 (18%) |
|  |  | 2 | 6905 (17%) | 72,171 (18%) |
|  |  | 3 | 8657 (22%) | 83,308 (21%) |
|  |  | 4 | 8214 (21%) | 78,074 (20%) |
|  |  | 5 (least deprived) | 9792 (25%) | 91,684 (23%) |
|  | **Year of birth** | <1934 | 2307 (6%) | 17,217 (4%) |
|  |  | 1934-1938 | 7239 (18%) | 55,199 (14%) |
|  |  | 1939-1943 | 10,787 (27%) | 86,002 (22%) |
|  |  | 1944-1948 | 12,216 (31%) | 126,071 (32%) |
|  |  | 1949-1953 | 7448 (19%) | 110,203 (28%) |
| **IQVIA** | **Sex** | Male | 4088 (39%) | 20,510 (47%) |
|  |  | Female | 6264 (61%) | 22,767 (53%) |
|  | **Insurance type** | Statutorily insured | 546 (5%) | 2692 (6%) |
|  |  | Statutorily insured family member | 56 (0.5%) | 294 (0.7%) |
|  |  | Statutorily insured pensioner | 8819 (85%) | 35,996 (83%) |
|  |  | Privately insured | 931 (9%) | 4295 (10%) |
|  | **Year of birth** | 1932 -1935 | 12,45 (12%) | 9070 (21%) |
|  |  | 1936 – 1939 | 1800 (17%) | 9166 (21%) |
|  |  | 1940 – 1944 | 2661 (26%) | 8457 (20%) |
|  |  | 1945 – 1948 | 2318 (22%) | 7294 (17%) |
|  |  | 1949 - 1952 | 2328 (22%) | 9290 (21%) |
| **DNR** | **Sex** | Male | 62,955 (37%) | 723,752 (47%) |
|  |  | Female | 107,037 (63%) | 818,359 (53%) |
|  | **Age (pyrs)** | 65-74 | 464,674 | 7,444,311 |
|  |  | 75-84 | 374,933 | 3,986,294 |
|  |  | 85-++ | 157,982 | 1,370,155 |
|  | **Calendar year (pyrs)** | 2000-2004 | 169,246 | 3,750,498 |
|  |  | 2005-2009 | 289,527 | 3,836,576 |
|  |  | 2010-2015 | 538,816 | 5,213,686 |
|  | **Civil status** | Married | 103,966 (61%) | 920,200 (60%) |
|  |  | Divorced | 20,301 (12%) | 179,138 (12%) |
|  |  | Widowed | 36,492 (22%) | 319,730 (21%) |
|  |  | Unmarried | 8663 (5%) | 98,423 (6%) |
|  |  | Unknown | 567 (0.3%) | 24,620 (2%) |
|  | **Highest attained educational level** | Low | 118,719 (70%) | 1,064,558 (69%) |
|  |  | Medium | 21,980 (13%) | 177,939 (12%) |
|  |  | High | 7257 (4%) | 54,959 (4%) |
|  |  | unknown | 22,033 (13%) | 244,655 (16%) |
|  | **CCI (pyrs)^[7]^** | 0 | 463,094 | 7,181,910 |
|  |  | 1 | 183,143 | 2,126,858 |
|  |  | 2 | 168,138 | 1,826,471 |
|  |  | 3 | 75,798 | 709,273 |
|  |  | 4+ | 107,416 | 956,247 |
| **eDRIS** |  |  |  |  |
|  | **Sex** | Male | 3409 (41%) | 168.206 (50%) |
|  |  | Female | 4885 (59%) | 166.137 (50%) |
|  | **SIMD^[8]^** | 1 (most deprived) | 1368 (17%) | 56.735 (17%) |
|  |  | 2 | 1592 (19%) | 65.033 (20%) |
|  |  | 3 | 1756 (21%) | 70.605 (21%) |
|  |  | 4 | 1853 (22%) | 72.600 (22%) |
|  |  | 5 (least deprvied) | 1720 (21%) | 68.777 (21%) |
|  | **Year of birth** | 1949 - 1951 | 6026 (73%) | 158.741 (47%) |
|  |  | 1952 - 1954 | 2268 (27%) | 175.602 (53%) |

| **Exposure** | | **Partially adjusted Hazard Ratio (95% CI)^[9]^** | | **Fully adjusted HR (95% CI)^1^** |
| --- | --- | --- | --- | --- |
| **Herpes^[10]^** | |  | |  |
|  | D-T- | 1 | | 1 |
|  | D+T- | 0.94 (0.88 to 1.02; p=0.127) | | 0.95 (0.88 to 1.02; p=0.177) |
|  | D+T+ | 0.91 (0.86 to 0.97; p=0.003) | | 0.91 (0.86 to 0.97; p=0.002) |
|  | D+T++ | 0.96 (0.80 to 1.13; p=0.604) | | 0.95 (0.80 to 1.12; p=0.528) |
|  | D+T+++ | 1.07 (0.86 to 1.33; p=0.571) | | 1.06 (0.85 to 1.32; p=0.601) |
| **Welsh Index of Multiple Deprivation (2011)** | | |  |  |
|  | 1 (most deprived) |  | | 1 |
|  | 2 |  | | 0.95 (0.90 to 0.99) |
|  | 3 |  | | 0.84 (0.80 to 0.88) |
|  | 4 |  | | 0.76 (0.72 to 0.8) |
|  | 5 (least deprived) |  | | 0.63 (0.6 to 0.66) |
| **Sex** | | | |  |
|  | Male |  | | 1 |
|  | Female |  | | 0.99 (0.96 to 1.02) |
| **Birth Year** | |  | |  |
|  | <1934 |  | | 1 |
|  | 1934-1938 |  | | 1.17 (1.12 to 1.24) |
|  | 1939-1943 |  | | 1.23 (1.16 to 1.30) |
|  | 1944-1948 |  | | 1.24 (1.16 to 1.34) |
|  | 1949-1953 |  | | 1.44 (1.29 to 1.62) |

| **Exposure** | | **Partially adjusted Hazard Ratio (95% CI)^[11]^** | | **Fully adjusted HR (95% CI)^1^** |
| --- | --- | --- | --- | --- |
| **Herpes^[12]^** | |  | |  |
|  | D-T- | 1 | | 1 |
|  | D+T- | 1.17 (1.08 to 1.27; p<0.001) | | 1.18 (1.09 to 1.28; p<0.001) |
|  | D+T+ | 1.06 (0.96 to 1.17; p=0.253) | | 1.08 (0.98 to 1.2; p=0.109) |
|  | D+T++ | 1.03 (0.80 to 1.33; p=0.834) | | 1.07 (0.83 to 1.38; p=0.603) |
|  | D+T+++ | 0.98 (0.69 to 1.40; p=0.918) | | 1.02 (0.71 to 1.45; p=0.929) |
| **Insurance type** | | |  | |
|  | Statutorily insured member |  | | 1 |
|  | Statutorily insured family member |  | | 0.74 (0.47 to 1.18) |
|  | Statutorily insured pensioner |  | | 1.01 (0.88 to 1.17) |
|  | Privately insured |  | | 0.43 (0.35 to 0.53) |
| **Sex** | | | |  |
|  | Male |  | | 1 |
|  | Female |  | | 1 (0.94 to 1.06) |
| **Birth Year** | |  | |  |
|  | 1932 – 1935 |  | | 1 |
|  | 1936 – 1939 |  | | 1.03 (0.95 to 1.11) |
|  | 1940 – 1944 |  | | 0.76 (0.69 to 0.84) |
|  | 1945 – 1948 |  | | 0.84 (0.73 to 0.95) |
|  | 1949 - 1952 |  | | 0.63 (0.52 to 0.77) |

| **Exposure^[13]^** | **Partially adjusted HR (95% CI)** | **Fully adjusted HR (95% CI)** |
| --- | --- | --- |
| D-T- | 1 | 1 |
| D+T+ | 0.94 (0.92 to 0.96; p<0.001) | 0.91 (0.89 to 0.93; p<0.001) |
| D+T++ | 0.98 (0.93 to 1.03; p=0.453) | 0.93 (0.88 to 0.98; p=0.008) |
| D+T+++ | 0.93 (0.88 to 1.00; p=0.033) | 0.89 (0.83 to 0.95; p<0.001) |
| **Sex** |  |  |
| Men | 1 | 1 |
| Women | 1.05 ( 1.04 to 1.06) | 1.06 ( 1.04 to 1.07) |
| **Age** |  |  |
| 65-74 | 0.10 ( 0.09 to 0.10) | 0.12 ( 0.12 to 0.12) |
| 75-84 | 0.46 ( 0.45 to 0.47) | 0.52 ( 0.51 to 0.52) |
| 85+ | 1 | 1 |
| **Calendar year** |  |  |
| 2000-2004 | 1.11 ( 1.10 to 1.13) | 1.11 ( 1.09 to 1.12) |
| 2005-2009 | 1.16 ( 1.15 to 1.18) | 1.16 ( 1.14 to 1.17) |
| 2010-2015 | 1 | 1 |
| **Civil status** |  |  |
| Single | 1 | 1 |
| Married | 0.91 ( 0.89 to 0.93) | 0.89 ( 0.87 to 0.91) |
| Divorced | 1.17 ( 1.13 to 1.20) | 1.11 ( 1.08 to 1.14) |
| Widowed | 0.97 ( 0.94 to 0.99) | 0.94 ( 0.91 to 0.96) |
| Unknown | 0.39 ( 0.36 to 0.42) | 0.47 ( 0.43 to 0.51) |
| **Education** |  |  |
| high | 0.91 ( 0.88 to 0.95) | 0.95 ( 0.92 to 0.99) |
| medium | 0.90 ( 0.88 to 0.92) | 0.92 ( 0.90 to 0.94) |
| low | 1 | 1 |
| unknown | 0.90 ( 0.89 to 0.92) | 0.94 ( 0.93 to 0.96) |
| **CCI^[14]^** |  |  |
| 0 | 1 | 1 |
| 1 | 1.58 ( 1.56 to 1.60) | 1.56 ( 1.54 to 1.58) |
| 2 | 1.42 ( 1.40 to 1.45) | 1.41 ( 1.38 to 1.43) |
| 3 | 1.76 ( 1.72 to 1.79) | 1.73 ( 1.70 to 1.77) |
| 4+ | 2.01 ( 1.98 to 2.05) | 1.98 ( 1.94 to 2.01) |

| **Exposure** | | **Partially adjusted Hazard Ratio (95% CI)^[15]^** | | **Fully adjusted HR (95% CI)^1^** |
| --- | --- | --- | --- | --- |
| **Herpes^[16]^** | |  | |  |
|  | D-T- | 1 | | 1 |
|  | D+T+ | 0.98 (0.64 to 1.50; p=0.923) | | 0.98 (0.64 to 1.49; p=0.911) |
|  | D+T++ | 1.85 (0.60 to 5.76; p=0.286) | | 1.89 (0.61 to 5.89; p=0.269) |
|  | D+T+++ | 2.17 (0.70 to 6.73; p=0.181) | | 2.18 (0.70 to 6.79; p=0.177) |
| **Scottish Index of Multiple Deprivation (2012)** | | |  |  |
|  | 1 (most deprived) |  | | 1 |
|  | 2 |  | | 0.78 (0.66 to 0.91) |
|  | 3 |  | | 0.61 (0.51 to 0.71) |
|  | 4 |  | | 0.59 (0.50 to 0.69) |
|  | 5 (least deprived) |  | | 0.45 (0.38 to 0.54) |
| **Sex** | | | |  |
|  | Male |  | | 1 |
|  | Female |  | | 1.04 (0.94 to 1.16) |
| **Birth Year** | |  | |  |
|  | <1952 |  | | 1 |
|  | >1951 |  | | 0.93 (0.80 to 1.08) |

1. Codes are displayed using notation from regular expression, where ^F0[0-3] stands for any code starting with F0, followed by either a 0, 1, 2 or 3 in third position, potentially followed by any other character or number. ^B0[02] stands for any coding starting with B0, followed by either a 0 or a 2 in third position.
2. Clinical terminology system used in GP practices in the UK
3. Anatomical Therapeutic Classification by the European Pharmaceutical Market Research Association (EphMRA) and the Pharmaceutical Business Intelligence and Research Group (PBIRG)
4. Anatomical Therapeutic Chemical (ATC) Classification by the WHO
5. British National Formulary
6. Welsh Index of multiple deprivation
7. Carlson Comorbidity Index
8. Scottish Index of multiple deprivation
9. Adjusted for random practice effect
10. Number of treatments: T+=treated once; T++=treated twice; T+++=treated thrice or more often
11. Adjusted for random practice effect
12. Number of treatments: T+=treated once; T++=treated twice; T+++=treated thrice or more often
13. Number of treatments: T+=treated once; T++=treated twice; T+++=treated thrice or more often
14. Carlson Comorbidity Index
15. Adjusted for random practice effect
16. Number of treatments: T+=treated once; T++=treated twice; T+++=treated thrice or more often
